# Targeted transcutaneous cervical spinal cord stimulation promotes upper limb recovery in spinal cord and peripheral nerve injury

**DOI:** 10.1101/2022.02.15.22269115

**Authors:** Santosh Chandrasekaran, Nikunj A Bhagat, Richard Ramdeo, Sadegh Ebrahimi, Pawan D Sharma, Adam Stein, Doug G Griffin, Susan J Harkema, Chad E Bouton

**Author notes:** **Corresponding authors:** Santosh Chandrasekaran, and Chad E Bouton.

## Abstract

Long-term recovery of limb function is a significant unmet need in people with paralysis. Targeted neuromodulation through epidural stimulation of the spinal cord, when paired with intense physical therapy, has shown promising results towards restoring volitional control of the upper limb in people with tetraplegia. Non-invasive neuromodulation of the cervical spinal cord using transcutaneous spinal cord stimulation (tSCS) has shown similar improvements in upper-limb motor control rehabilitation. However, the potential rehabilitative effects of activating specific cervical spinal segments in a targeted fashion using tSCS has largely remained unexplored. We show in two individuals with SCI that tSCS of the cervical spinal cord resulted in up to 1136% increase in exerted force, with minimal physical therapy. We also show, for the first time, the effectiveness of targeted tSCS in restoring strength (407% increase) and dexterity to the digits in an individual with paralysis of the hand due to a peripheral nerve injury. Furthermore, we believe this is the first study to document a 3-point improvement in sensation in people with SCI or peripheral injury after receiving tSCS. Lastly, participant gains persisted after a one-month period void of stimulation, suggesting tSCS may lead to lasting benefits. Non-invasive targeted spinal cord stimulation shows tremendous promise as a safe and effective therapeutic approach with broad applications.

## Introduction

Spinal cord and peripheral nerve injury can result in paralysis due to the disruption in the transmission of neural signals. More than half of spinal cord injuries (SCIs) occur at the cervical level (NSCISC and University of Alabama at Birmingham, 2021), and regaining voluntary control of the hand and arm is the highest priority in such cases (Anderson, 2004). However, the chances of regaining hand and arm function are exceedingly low beyond 12-18 months post injury (Fawcett et al., 2006).

Electrical stimulation of the lumbar spinal cord using epidurally placed electrodes has recently shown great promise in evoking voluntary movements of the lower limb during stimulation after SCI, in rats (van den Brand et al., 2012), non-human primates (Capogrosso et al., 2016) and humans (Harkema et al., 2011; Angeli et al., 2014, 2018; Gill et al., 2018; Wagner et al., 2018). Additionally, when paired with intense motor training, lumbar epidural stimulation has also demonstrated persistent increase in voluntary control of the lower limbs even in the absence of stimulation (Rejc et al., 2017; Angeli et al., 2018; Wagner et al., 2018; Seáñez and Capogrosso, 2021; Rowald et al., 2022). Epidural stimulation has been shown to primarily engage the large-to-medium size sensory afferent fibers present in the dorsal column and roots of the spinal cord (Rattay et al., 2000). Local spinal circuits constitute of these afferent fibers forming synaptic connections with spinal interneurons and motoneurons. Pharmacological and computational experiments (Capogrosso et al., 2013) suggest that activation of the dorsal fibers increases the excitability of the local spinal circuitry including the efferent fibers. Expanding this idea further, recent studies have demonstrated that epidural stimulation targeting cervical spinal cord could activate (Greiner et al., 2020) and restore upper limb movement (Lu et al., 2016). Though extremely promising, the invasive nature of epidural stimulation is a major hurdle for clinical translation and long-term rehabilitation (James et al., 2018; Taccola et al., 2020).

Meanwhile, non-invasive, persistent stimulation of the spinal cord using transcutaneous electrodes has been demonstrated to evoke voluntary movements in both upper (Freyvert et al., 2018; Gad et al., 2018; Inanici et al., 2021) and lower limbs (Sayenko et al., 2015, 2019). Moreover, recent evidence suggests that transcutaneous spinal cord stimulation (tSCS), like epidural stimulation, exerts its neuromodulatory effect on motoneurons trans-synaptically via activation of large-to-medium size sensory afferent fibers (Ladenbauer et al., 2010; Hofstoetter et al., 2018) and can also improve descending supraspinal drive (Guiho et al., 2021). Only recently, has it been shown that pairing tSCS at the cervical levels with intense motor training can result in sustained improvements in hand and arm function (Benavides et al., 2020; Zhang et al., 2020; Inanici et al., 2021). Taken together, tSCS can be a promising rehabilitative tool for people with SCI.

The commercially available electrodes employed for tSCS, however, are relatively large resulting in a wider distribution of the applied current. Additionally, being placed over the skin, the electrodes are also distant from the dorsal fibers of the spinal cord with layers of intervening connective and bony tissue including the dorsal aspects of the cervical vertebrae. These factors may limit the effectiveness and specificity of tSCS for the recruitment of target motoneuron pools which could be a major drawback since epidural stimulation studies have shown the importance of targeted stimulation (Wagner et al., 2018) in restoring voluntary muscle control.

We performed targeted transcutaneous stimulation of the cervical spinal cord paired with minimal physical therapy in two individuals classified as having a motor complete SCI and one individual with a peripheral nerve injury. Stimulation was targeted to specific cervical levels using a custom electronically configurable electrode array, with each electrode having a small form factor (10 mm x 10 mm). Modulating the spatial configuration of the stimulation electronically allowed us to extensively map the recruitment profile of the upper-limb muscles based on the location of stimulation. This enabled us to choose the precise location of stimulation to achieve maximal recruitment of the muscle group of interest. Even though participants received stimulation and motor training only once per week, we observed a rapid increase in both volitionally controlled muscle activity and effective force within a period of 5-6 weeks. However, the observed gains were restricted to muscles that generated at least a measurable amount of force at the beginning of the intervention. In the subject with peripheral nerve injury, we observed a rapid increase in functionality of the affected digits.

Taken together, this study demonstrates the advantages of using a highly configurable electrode array in conjunction with minimal physical therapy in restoring volitional control of upper-limb movement using tSCS. Also, this is the first study to demonstrate the benefits of tSCS for improving hand function in a case of peripheral injury.

## Methods

### Participants

We performed transcutaneous stimulation of the cervical spinal cord in two individuals with tetraplegia resulting from C5 level motor complete spinal cord injuries and one individual with paralysis of the left hand due to peripheral nerve injury (median nerve at the wrist and suspected brachial plexus injury), but with an otherwise intact spinal cord. The details of the participants are summarized in Table 1. All procedures were approved by the Northwell Health Institutional Review Board. The study has been registered with ClinicalTrials.gov (NCT04755699).

**Table 1.** Study participants information. Demographic and injury-related information for each participant.

| Participant | Age Range | Gender | Injury type | Time since injury (yrs.) |
| --- | --- | --- | --- | --- |
| CTS01 | late 40s | F | Median nerve, Brachial plexus | 4 |
| CTS02 | early 20s | M | SCI: C5, ASIA A | 2 |
| CTS03 | mid 30s | M | SCI: C5, ASIA B | 7 |

### Experimental sessions

Participants visited the lab for their intervention sessions once per week. Their heart rate and blood pressure were measured at the beginning and end of each session. Baseline GRASSP assessments were carried out in two initial sessions, scheduled a month apart, to establish an initial baseline. For finer measurements of voluntary muscle control, we measured both the evoked force and electromyographic (EMG) activity in the muscles of the hand and arm. For efficiency, we restricted our measurements to only one side for all participants. For participants with SCI, we chose the left side. For the participant with the peripheral nerve injury, we chose the side that was affected by the resultant paralysis, i.e., the left side. Bipolar surface EMG electrodes were used to record muscle activity from the following muscles of the left arm and hand: biceps brachii (BIC), triceps brachii (TRI), flexor digitorum profundus (FDP), extensor digitorum communis (EDC) and abductor policis brevis (ABP). Force generated by the isometric flexion of the muscles of interest and EMG activity were simultaneously recorded at the start of each session.

For evaluating the forces and EMG activity generated by the tricep muscle, as in the case of participants CTS02 and CTS03, the participants’ arms were extended out in front of them, with the hand in a neutral position and a load cell placed under the ulnar head prominence at the wrist (Figure 1C). The participants were instructed pushed down on to the load cell in this position. The elbow was resting on the table and prevented from lifting during the task. For evaluation of the D1 interphalangeal (D1 IP) joint and lateral pinch, as in the case of participant CTS01, a custom rig was used as shown in Figure 1D-E. The participant was asked to push down on the sensor using their thumb. For all force measuring tasks, the experimenter cued the participants to push against the force sensor for 3-5 secs and then to relax for 3-5 secs. The task was performed in 2 sets of 5 trials each with a 60-90 secs of rest period between the sets. Verbal encouragement was provided to encourage the participants to generate maximal maintainable force.

**Figure 1.**
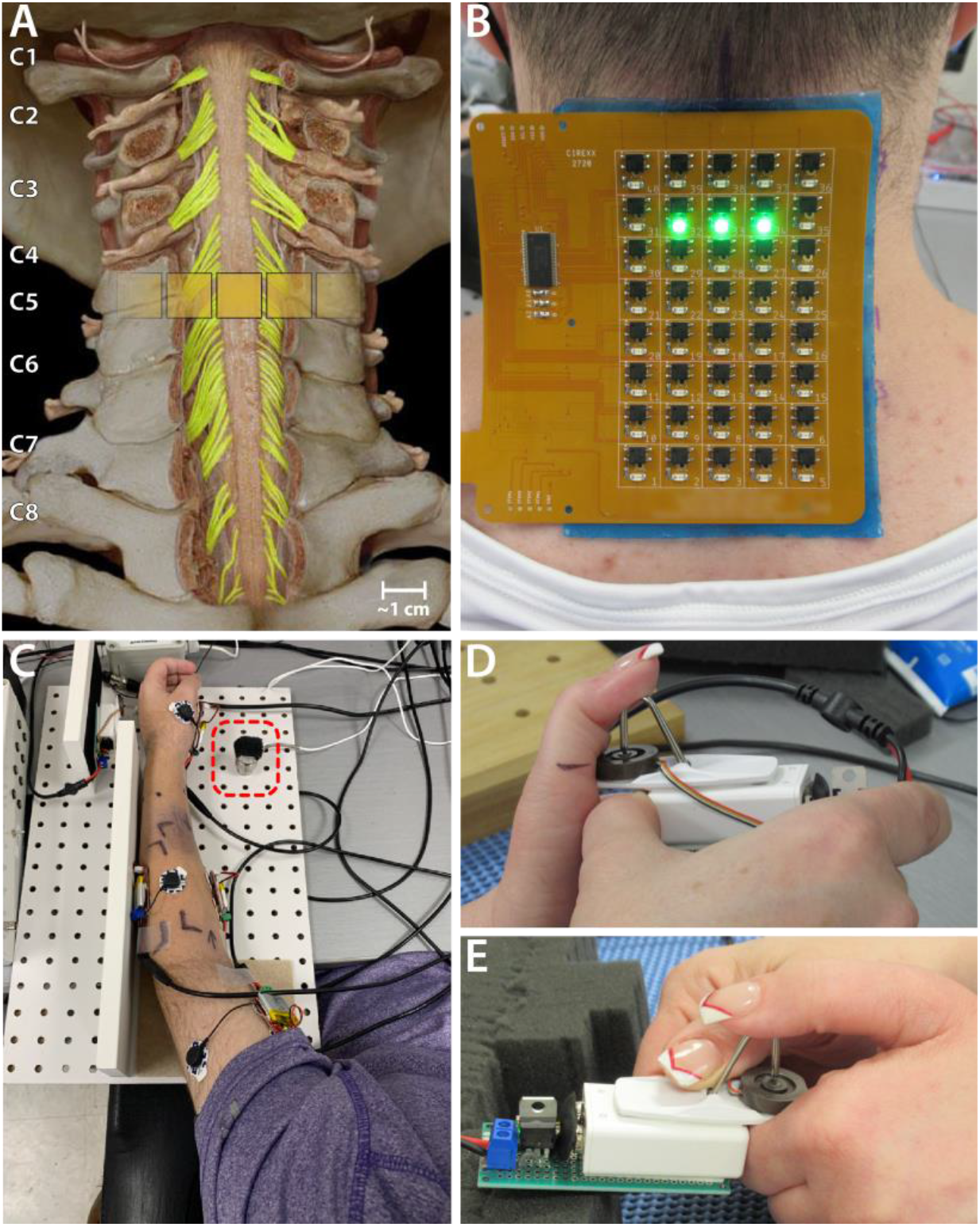
Experiment setup. **A)** Schematic showing the location of a 1 × 3 activated electrode configuration superimposed over the human spinal cord showing the dorsal column and roots. **B)** The custom electronically configurable electrode array placed over the cervical spinal cord of a study participant with a 1 × 3 configuration of activated electrodes (green LEDs). **C)** Setup for measuring tricep force. The ulnar protrusion of the wrist is placed over the 25 lb load cell during the task (red dashed square). **D-E)** Using the custom-built rig with a 10 lb load cell (gray puck) for measuring the force using D1 interphalangeal joint and lateral pinch, respectively in case of participant CTS01.

While administering tSCS as a therapeutic intervention, the participant performed the same tasks as described above. This served the dual purpose of providing physical therapy while receiving tSCS as well as measuring the evoked volitional EMG activity and force.

### Transcutaneous spinal cord stimulation (tSCS)

Transcutaneous spinal cord stimulation was provided using a custom-built stimulator and electrode array. The flexible PCB electrode array consisted of electroless nickel immersion gold (ENIG) or immersion silver-plated square contacts (10 mm x 10 mm) arranged in an 8 × 5 pattern with a 1 mm inter-electrode separation. Electrical stimulation to any combination of the 40 electrodes within the array, was controlled using solid-state relays mounted above each of the electrodes and a custom MATLAB-based GUI. Miniature green LEDs mounted on the dorsal surface of the electrode array, visually indicated the active electrodes. To target specific cervical segments (Figure 1A), an electrode configuration of 1 × 3 (Figure 1B) was used wherein 3 adjacent contacts within a single row, spanning the midline, were used. The electrode array was affixed to the back of the neck using a rectangular piece of proprietary hydrogel. To ensure consistency in placement of the array between sessions, we used the inion of the external occipital protuberance as a landmark. Distances measured from the inion were used to place the electrode array and identify the location of stimulation. Two 5 × 10 cm rectangular self-adhesive hydrogel electrodes (Axelgaard Manufacturing Co., Ltd., USA) placed along the midline over the lumbar spinal cord served as return electrodes (anodes).

Stimulation consisted of a 10 kHz multiphasic sinusoidal waveform with pulse duration of 1 ms for generating recruitment profiles. The waveform length was reduced to 0.5 ms during therapy to reduce neck muscle activation and increase participant comfort. Similar pulse duration has been used in previously published work (Alam et al., 2020; Benavides et. al, 2020). The stimulation frequency was chosen to be 3 Hz for characterizing recruitment curves and 50 Hz during therapy.

While administering tSCS as a therapeutic intervention, the stimulation amplitude was chosen to be 140-160 mA. This amplitude was chosen based on patient comfort but at the same time was an amplitude that did not interfere with the tasks being performed by the participant. Stimulation was delivered for 45-60 minutes and was paired with the participant performing the force measurement tasks described above.

After 16 weeks of receiving of tSCS, we administered a 3-week period when the participant received no stimulation at all. They continued to perform the tasks that constituted their physical therapy as described above. Weekly administration of tSCS was resumed after this ‘No Stim’ period.

### EMG signal processing

We simultaneously recorded bipolar EMG from the following muscles of the left arm and hand: biceps brachii (BIC), triceps brachii (TRI), flexor digitorum profundus (FDP), extensor digitorum communis (EDC) and abductor policis brevis (ABP). We used pre-gelled Ag/AgCl electrodes, a differential amplifier (AD6221) with a bandpass filter (20-498Hz), and a signal digitizer (PicoScope® Model 4824A). The sampling rate used was 10 MHz while characterizing the recruitment profile of the upper limb muscles and 10-20 kHz during task performance. For characterizing the area-under-the curve (AUC), the EMG signal was filtered using a 60Hz IIR comb filter and Butterworth bandpass filter between 10-1000 Hz using a digital filter in MATLAB.

### Recruitment profile

Stimulation consisted of a 1ms-long multiphasic pulse of the 10 kHz sinusoidal waveform at a stimulation frequency of 3 Hz delivered using an electronically selected configuration of three contacts within a single row spanning the midline. Stimulation amplitudes tested ranged from 100 mA to up to 225 mA in intervals of approximately 25 mA. This was repeated for each of the eight rows of electrodes on the array. The Picoscope 6 acquisition software (Pico Technology, Cambridgeshire, UK) was used to trigger acquisition of a 100 ms-long EMG signal following each stimulation pulse. For each stimulation amplitude and at each electrode row, we recorded such an EMG signal from 20-30 repetitions of the stimulation pulse. All the data was imported into MATLAB for further analysis. From each of the EMG signals recorded, we isolated a snippet starting from 5 ms and ending at 55ms after the stimulation artifact. We measured the peak-to-peak amplitude (P2P) for this snippet. It was included for analysis only if the maximum amplitude of the snippet was greater than 5 times the standard deviation of baseline signal of that recording channel. For each muscle, the P2P amplitudes were normalized to the maximal P2P amplitude recorded across all amplitudes and electrode rows.

We also measured the stimulation current of maximal activation for each muscle. For this, at each cervical level, we increased the amplitude until the reflexive activation of the muscle was visually determined to be consistent for every stimulation pulse and did not change for any small increase in amplitude. This stimulation amplitude was noted as the Maximal Activation Current for that muscle at the cervical level. This procedure was repeated for each muscle.

### Functional Outcome

In addition to the areas recommended in the GRASSP assessment for sensibility evaluation, we tested few additional relevant spots on the dorsal and volar aspect of the hand to obtain a comprehensive profiling of the sensory changes secondary to the intervention sessions. To provide encouragement and also observe the true capability of the participant, we initially let the GRASSP motor assessment tasks run for more than the stipulated time of 75 secs.

### Statistical Analysis

For statistical analysis of the evoked force, we compared the forces recorded in three stages of the study (start, no-stim and end of study). For the early stage, we chose the first three consecutive sessions after start of weekly tSCS sessions. For the no-stim stage, we chose three sessions during the period when tSCS was not being delivered. For the end of study stage, we chose the last three consecutive sessions. For the force measurements of the tricep muscle in case of the participants with SCI, we performed a repeated-measures Analysis of Variance (ANOVA) between the average values of force obtained in the three epochs. For the force measurements involving the D1 IP muscle in case of the participant with the peripheral nerve injury, we performed a t-test. We employed the Bonferroni correction for all multiple comparisons.

## Results

Our results demonstrate that transcutaneous spinal cord stimulation can be used to activate specific motoneuron pools in the cervical spinal cord. Moreover, targeted transcutaneous spinal cord stimulation paired with minimal physical therapy resulted in a substantial and sustained increase in muscle activity and strength in specific upper-limb muscles in two patients with motor complete cervical SCI and one participant with a peripheral injury. All three participants received stimulation for up to 1hr, once per week. Blood pressure and heart rate monitoring at the start and end of each session showed no adverse effects of the stimulation.

### Recruitment of upper limb muscles through targeted tSCS

We characterized the recruitment profile of the different motor pools innervating the upper limb muscles. To determine the effect of the location of stimulation along the rostro-caudal axis on the recruitment of the upper limb muscles, we sequentially delivered stimulation to different electrode triplets in the array (Figure 2A-C); also see Supplementary Figure 1 for an example stimulation-evoked EMG activity. Simultaneously, we recorded EMG activity from 5 muscles of the left arm and hand. Stimulation amplitude was increased and the corresponding increase in EMG amplitude was used as a measure of recruitment. We hypothesized that the recruitment pattern through transcutaneous stimulation would reflect the rostrocaudal segment-wise distribution of the upper limb motor nuclei in the cervical spinal cord. The recruitment profiles showed a distinct and consistent shape in all participants. Almost all the muscles exhibited the best recruitment when stimulation was delivered around the C5-C6 level. Stimulating using rostral electrodes primarily activated the biceps (Figure 2D, G and E, H), except in CTS03. Activation of the triceps invariably was strongest when stimulation was localized around the C5-C6 level. Stimulation using electrodes positioned more caudally resulted in decreased recruitment across all muscles. C8-T1 level stimulation showed recruitment of FDP and APB in participant CTS03 (Figure 2C, F and I). In participant CTS01, APB showed only weak recruitment irrespective of the location of stimulation, possibly due to the suspected brachial plexus injury. The fact that two participants had a spinal cord injury and metallic implants in the cervical vertebrae did not result in an obvious difference in the recruitment profile of the upper-limb motor pools is noteworthy. Additionally, we also determined the current required for maximal activation (Max. Activation Current) for each of the upper limb muscles. The thresholds of activation showed similar results as the recruitment profiles with biceps and/or triceps muscles having a low threshold of activation during rostral stimulation (Figure 2G, H and I).

**Figure 2.**
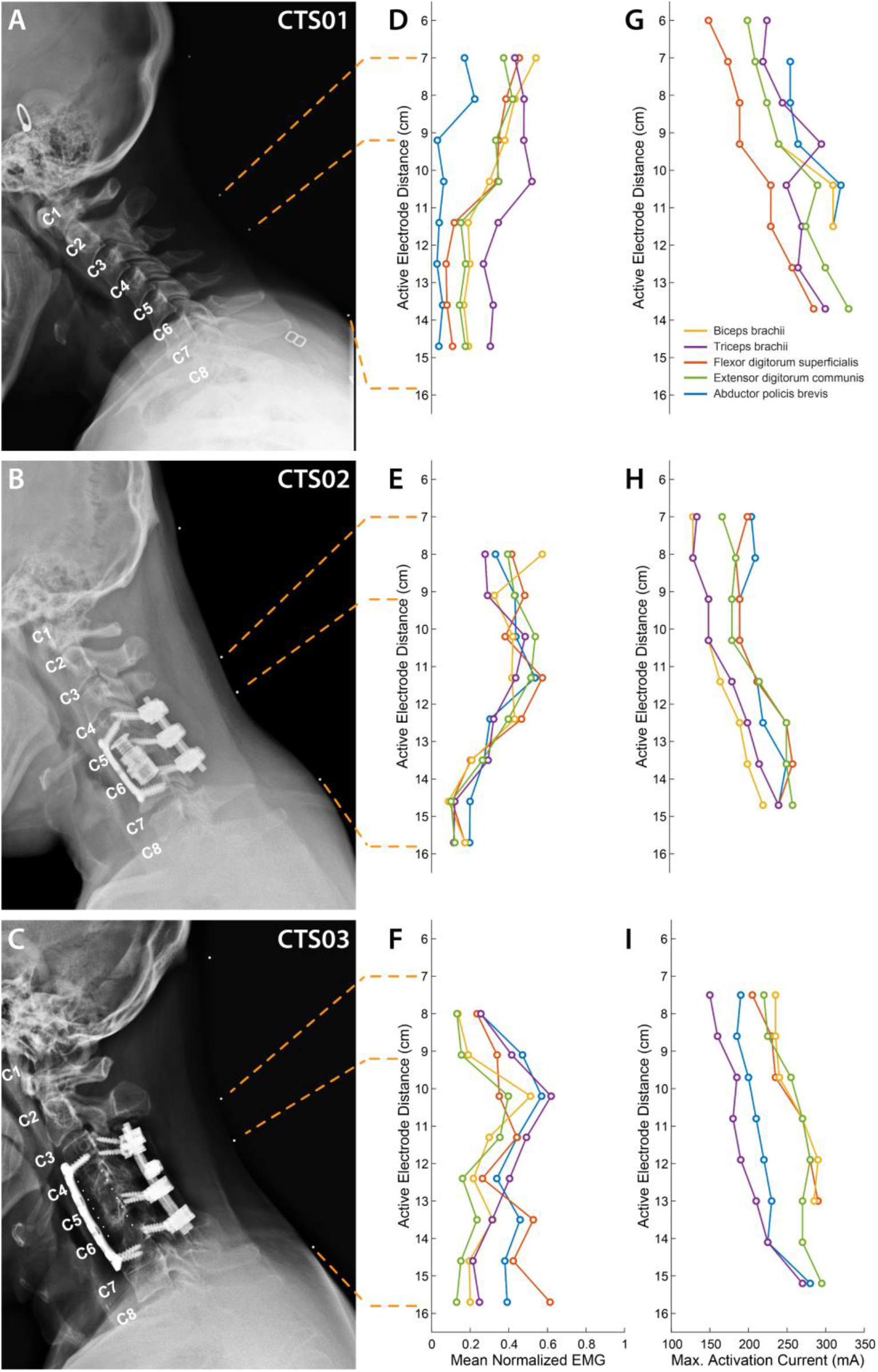
Muscle recruitment profile during cervical tSCS. **A-C)** X-ray images in the sagittal plane with radio-opaque markers on the neck (white dots) for the three participants. The topmost marker identifies the inion of the external occipital protuberance. The second and third markers identify points 7 cm and 9.2 cm respectively, from the inion signifying the first and third rows of a putative electrode array whose first row of electrodes was aligned at 7cm from the inion. The last marker identifies the location of the last row of the putative electrode array at 15.7 cm from the inion. The cervical labels mark the exit point of the respective dorsal roots. **D-F)** Mean activation of the 5 muscles across all stimulation amplitudes mediated by tSCS through each of the 8 electrode rows. **G-I)** Stimulation amplitude that resulted in maximal activation of each of the 5 muscles.

### Increased muscle activity and force generated with tSCS

Previous spinal cord stimulation studies have generally shown improvements in muscles that have at least some residual activity. Given this, we focused on muscles with at least some residual activity in our participants. Since both SCI participants exhibited no movement in their fingers, we focused on the volitional control of their tricep muscle. Meanwhile, as the participant with the brachial plexus injury (CTS01) had residual movement in the fingers of the left hand, we focused on the volitional control of the D1 interphalangeal (D1 IP) joint and lateral pinch. Here we report the forces and EMG recorded from these specific muscles before any tSCS was delivered during the session. We chose the site of stimulation based on the choice of muscle and its activation profile. For the participant with the brachial plexus injury, we focused on thumb muscles, and delivered tSCS targeted at the C5 level (about 8 cm from the inion of the external occipital protuberance) based on the activation profile for the APB muscle (see Figure 2A, D and G). For participants CTS02 and CTS03, we delivered tSCS targeted at the C6 level (about 10 cm from the inion of the external occipital protuberance).

We observed an average increase of 407% (t-statistic = -5.26, p < 0.05) in the force generated by the flexion of D1 IP joint (Figure 3A) in the ‘No Stim’ period at the end of 16 sessions compared to the forces at the start of the study. This reduced a little to 287% increase by the end of the study (30 sessions). This was accompanied by an increase in EMG activity of the left APB muscle (Figure 3B). We did not observe any major increase in the lateral pinch force during this period. We observed an average increase of force generated by the left tricep muscle of up to 893% (at the end of 17 sessions) and 825% (at the end of 16 sessions) for CTS02 and CTS03, respectively, in the ‘No Stim’ period compared to the start of the study. This increased to 1136% (29 sessions) and 1035% (31 sessions), respectively (F = 13.99, p < 0.01), (Figure 3C, Supp. Fig. 1). We also observed a corresponding increase in EMG activity for the tricep muscle (Figure 3D).

**Figure 3.**
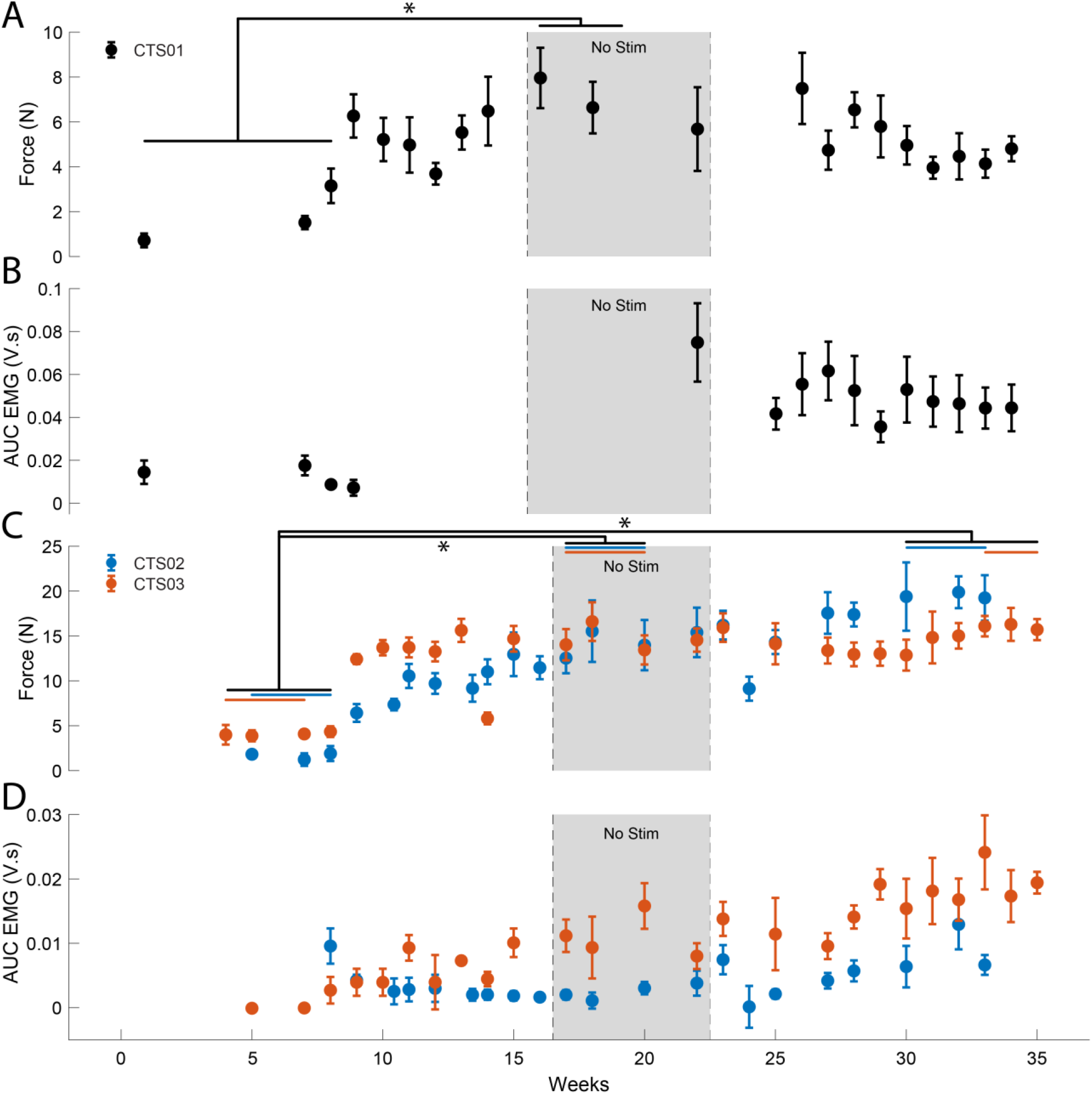
Increase in muscle activity and force after tSCS. **A)** The force generated by the left D1 IP joint in case of participant CTS01. * indicates a significant difference (Two-sample t-test with Bonferroni correction for multiple comparisons) **B)** The AUC of the EMG activity recorded from the left APB muscle during the task of force generation using the D1 IP joint. **C)** The force generated by the left tricep muscle in case of participant CTS02 (blue) and CTS03 (orange). * indicates significant difference (Repeated measures ANOVA and Bonferroni correction for multiple comparison). The blue (CTS02) and orange (CTS03) bars indicate the specific data points from each participant that were included for the statistical analysis. **D)** The AUC of the EMG activity recorded from the left tricep muscle during the task of force generation using the left tricep muscle.

### GRASSP assessments

To evaluate the clinical significance of the progress showed by each participant, we also performed standard clinical assessments, namely the GRASSP test. The GRASSP strength test showed only minor increases in the movements assessed, especially in the participants with SCI including the tricep muscle which was specifically targeted in this study (Figure 4A). GRASSP sensibility test showed decreases in sensation threshold at many locations in the hand in all participants (Figure 4B). For the participant with the brachial plexus injury, most of the improvement was observed in the thumb as well as the ulnar side of the hand. GRASSP strength showed increases in D1 IP and D5 abduction movements (Figure 4A). GRASSP sensibility showed up to a 3-point increase in sensation at the pinky tip (Figure 4B).

**Figure 4.**
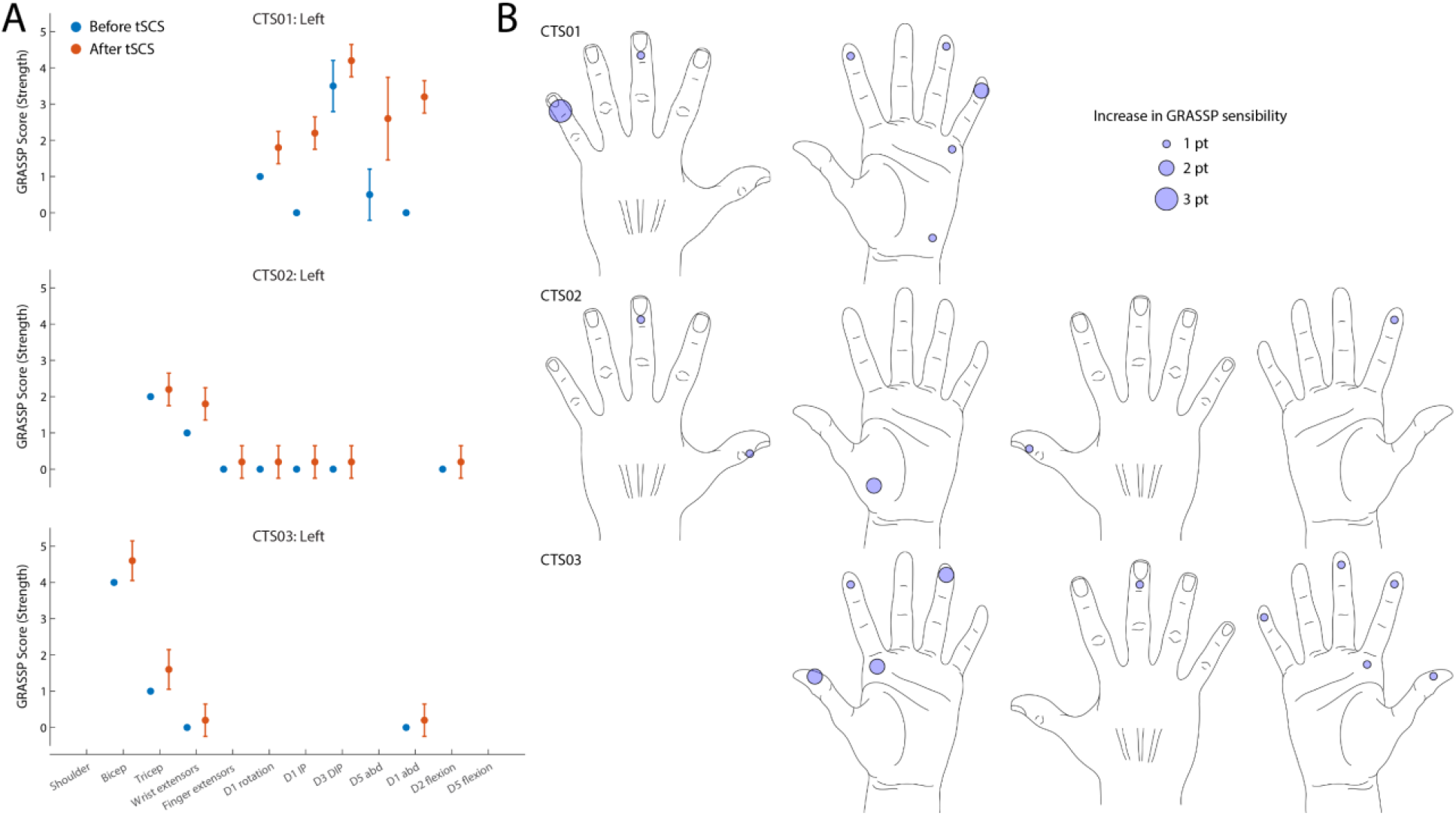
GRASSP assessments. **A)** GRASSP strength change from baseline **B)** GRASSP sensibility change from baseline. Circles show 1- to 3-point increase in sensory perception as defined by the GRASSP sensibility scale. **C)** GRASSP prehension change in nut threading task for CTS01.

The nut threading task included in the GRASSP prehension tests showed an improvement in performing the right grasp (Figure 5A) and a decrease in the time taken to perform the task (Figure 5B). Participant CTS01 also showed improvement in range of movement of thumb abduction, flexion, and extension as well as index abduction (Supp. Fig. 2). Participants also shared their anecdotal descriptions of the effect of receiving tSCS. These described increased control in moving their arms and being able to give “stronger hugs.”

**Figure 5.**
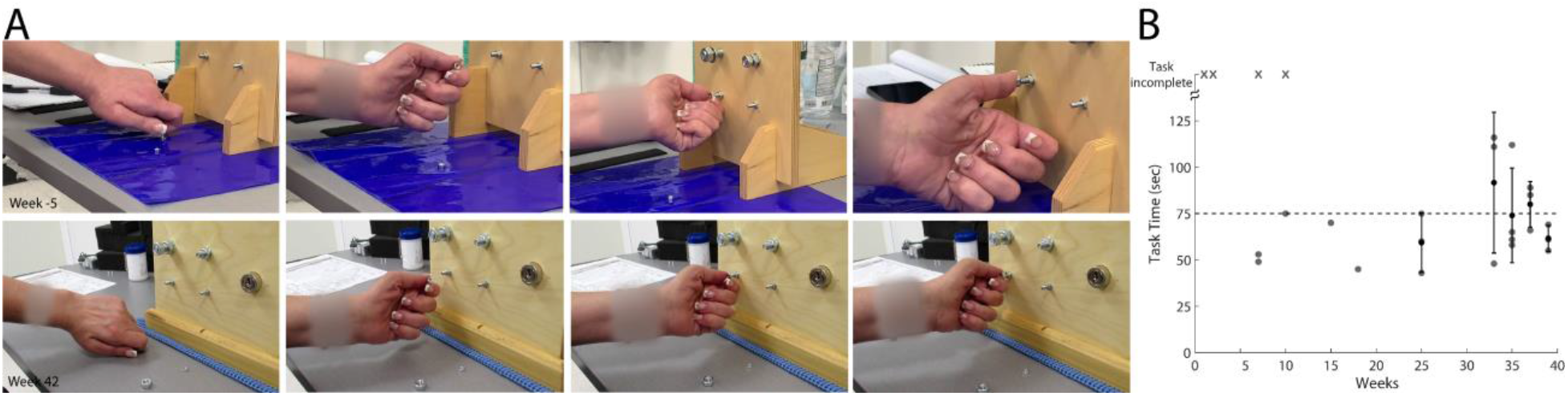
GRASSP Prehension Test – Nut threading. **A)** Snapshots showing improvement in grasping behavior from before tSCS therapy (first row) and week 42 of therapy (second row). **B)** Scatter plot shows the improvement in the time required to perform the task. ‘x’ denotes that the participant never managed to complete the task.

## Discussion

In this study, we observed a substantial increase in volitionally generated force and EMG activity in specific muscles after paralysis from tSCS in two individuals with SCI and one with a peripheral injury (median nerve and suspected brachial plexus injury). All three participants showed these changes within a brief period of receiving tSCS. Interestingly, the improvements were restricted to the muscles that showed at least some measurable amount of force at the beginning of therapy. For instance, neither of the participants with SCI registered any force during finger flexion. We did not observe any improvement in the forces and EMG activity generated during finger flexion in either participant with SCI even when the activity was performed over many weeks while receiving tSCS. This could be due to non-optimal tSCS patterns or that the severity of the injuries was too severe to benefit from this type of therapeutic intervention.

Our results are comparable to those demonstrated by other studies involving cervical tSCS in people with SCI (Inanici et al., 2021). However, our design involved participants receiving spinal cord stimulation and minimal physical therapy for 1-2 hrs only once every week. Importantly, our stimulation was targeted to achieve maximal recruitment of the muscle group of interest. This suggests that targeted tSCS could improve the efficacy of spinal cord activation and achieve restoration of volitional control even with minimal physical therapy.

The electrode configuration we used in this study was a 1 row x 3 columns configuration spanning the spinal cord midline. The effectiveness of such an electrode configuration in recruitment of specific motorneuron pools has been demonstrated in able-bodied individuals (Gerasimenko et al., 2015; Krenn et al., 2015). It is highly likely that this configuration resulted in the stimulation of the dorsal column fibers thereby activating motor nuclei in off-target cervical levels and thus reducing specificity. Activation of the dorsal roots via lateralized stimulation could result in greater selectivity in the motoneuron pools being activated. However, an earlier study with lateralized stimulation showed an increase in side-specific activation and not across spinal levels (Calvert et al., 2019). Previous studies have explored the idea of targeting stimulation above and below the injury level with the aim of enhancing the activity of the descending inputs as well as local circuitry below the site of injury (Zhang et al., 2020; Inanici et al., 2021). With a high cervical location of SCI (C5) for the participants in this study, most of the stimulation was restricted to being targeted at or below the injury level (Freyvert et al., 2018). In fact, stimulation targeted at higher cervical levels was perceived as uncomfortable by the participants. This demonstrates that tSCS can be tailored to suit patient comfort, target only those cervical levels innervating the muscle of interest, and still result in significant motor improvements.

Furthermore, we used a custom electrode array that could be electronically configured. This allowed efficient mapping and can support dynamic spatial pattern switching. In future studies, this feature can be combined with brain-computer interfaces (BCIs) to switch spatial patterns based on movement-related information decoded from intracortical activity (Bouton et al., 2016; Sharma et al., 2016; Friedenberg et al., 2017; Chandrasekaran et al., 2021b). Such automated configuration of targeted tSCS based on user movement intentions could improve the usability of this technology by people with SCI while performing activities of daily living.

It was interesting to observe improvement in somatosensory perception in regions innervated by spinal roots from below the injury level in both participants with SCI. To our knowledge, this is the first study to document improvements in sensation in people with SCI or peripheral injury after receiving tSCS. Stimulating dorsal roots of the spinal cord has been demonstrated to relay somatotopically relevant sensory information (Chandrasekaran et al., 2020). Meanwhile, restoring somatosensation through intracortical stimulation has been repeatedly demonstrated in humans as well (Flesher et al., 2016; Chandrasekaran et al., 2021a; Fifer et al., 2021). It may be worthwhile to explore the benefits of pairing intracortical stimulation and tSCS for the long-term rehabilitation of somatosensation in SCI.

We believe this is the first study that performed tSCS in an individual with peripheral injury (Lopez and Sdrulla, 2021) that resulted in the rehabilitation of volitional control of hand movement. The rapid and significant improvement observed in the hand function in this case signifies major implications for a broader range of conditions affecting the central and peripheral nervous systems, including traumatic injuries and stroke.

## Data Availability

All data produced in the present study are available upon reasonable request to the authors.

## Acknowledgements

We would like to thank our participants for their extraordinary commitment to this study, their patience with the experiments, and the deep insights provided by them, the clinicians, and researchers at Feinstein Institutes for Medical Research. We would like to extend our deepest gratitude to Dan Sciubba, MD, David Bonda, MD and Karen Black, MD at Northwell Health for useful discussions and assistance related to the imaging sessions.

## Author Contributions

SC, NAB and CEB designed the study. SC, NAB, RR, SE and CEB performed the experiments and SC analyzed the data. NAB designed and tested the electrode array, with guidance from CEB. DGG performed an independent assessment of the motor and sensory capacity of the participants. PDS and SJH provided critical feedback and input. All authors contributed towards interpreting the results of the experiments. SC and CEB finished the initial draft of the manuscript and all authors provided critical review, edits, and approval of the final manuscript.

## Supplementary Figures

**Supplementary Figure 1.**
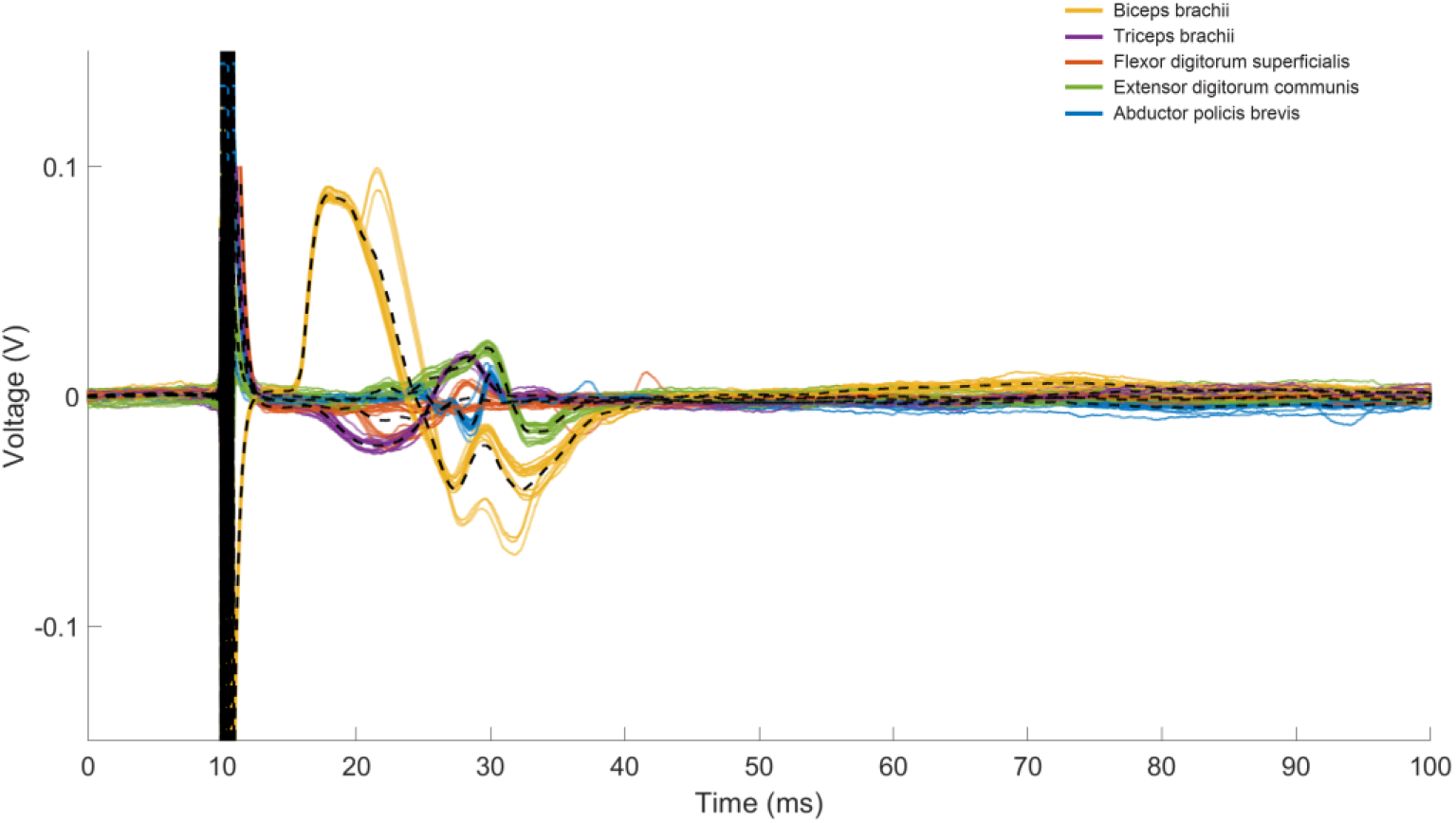
Example trace of muscle recruitment profile of the 5 muscles upon tSCS stimulation. This was for participant CTS02 when stimulated at 10.2 cm at 150 mA. Colored traces indicate the reflex activity evoked in each of the 5 muscles as shown in the legend. Dashed black line shows average of all the trials.

**Supplementary Figure 2.**
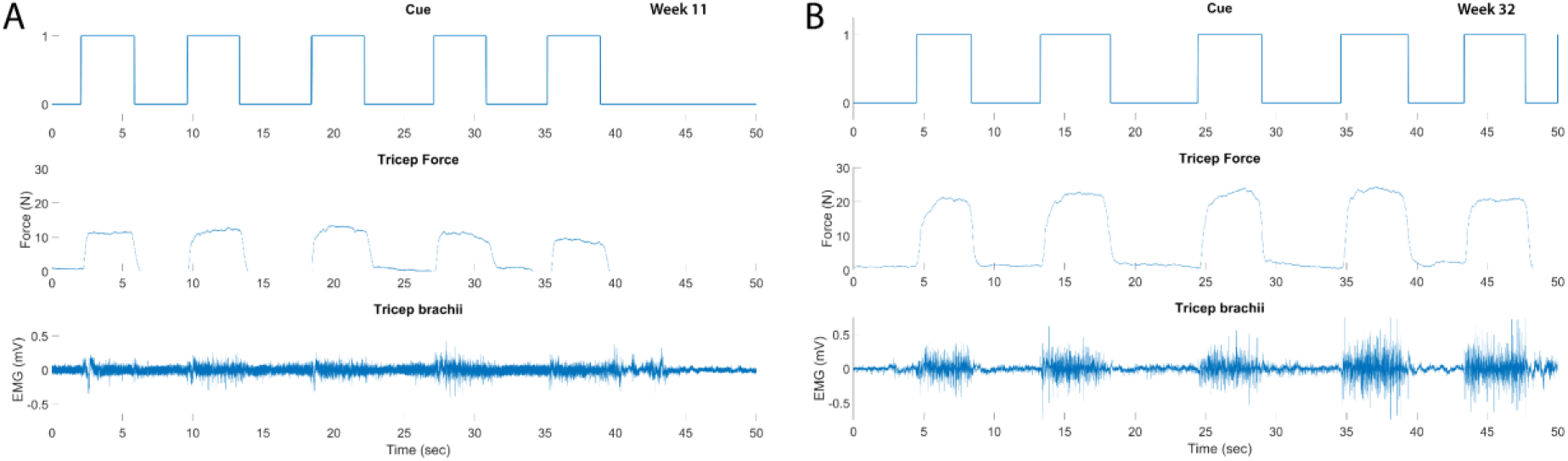
Example traces of cue, force measured during tricep extension and EMG recorded from triceps brachii for participant CTS02 from A) week 11 and B) week 32.

**Supplementary Figure 3.**
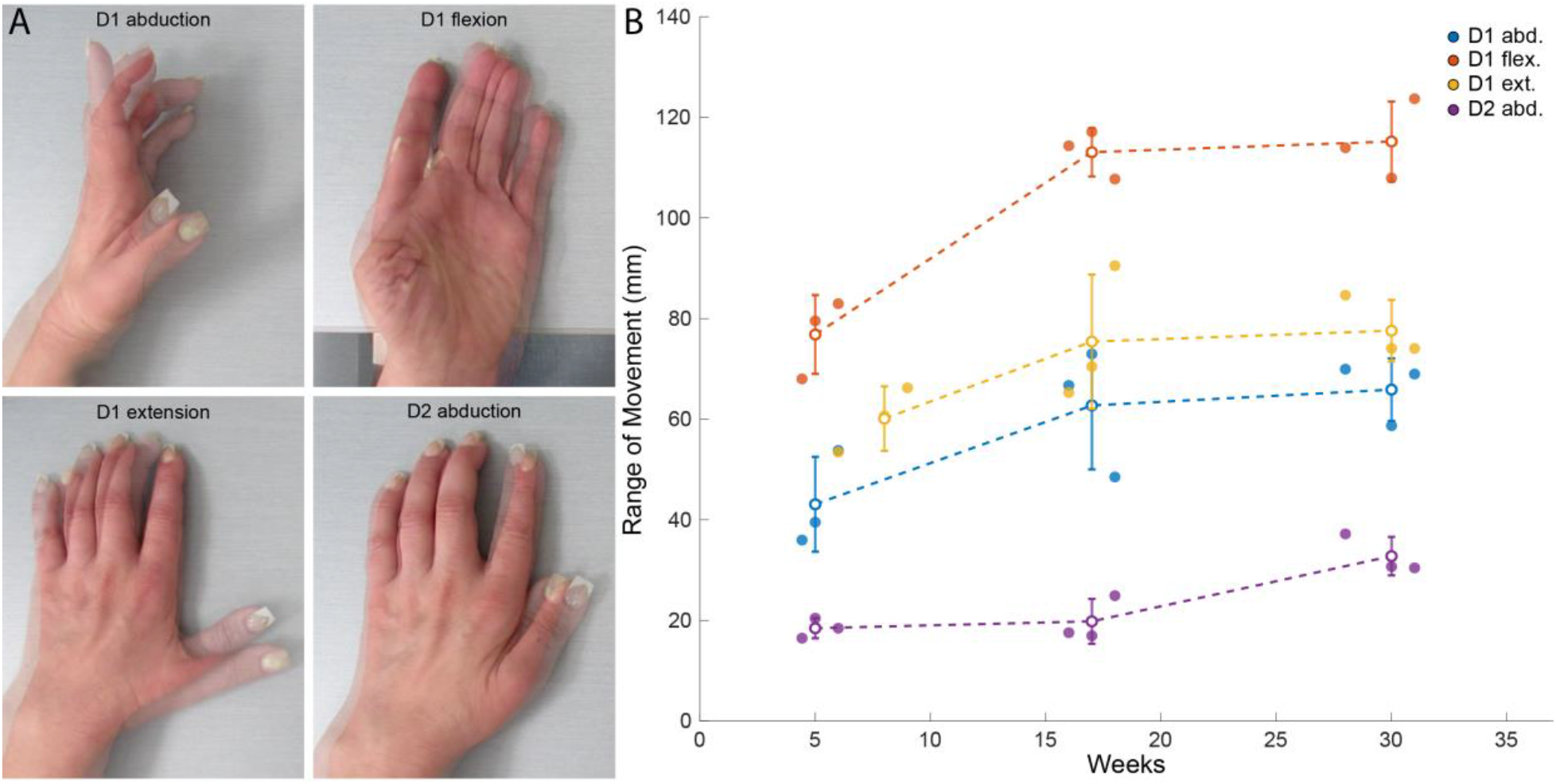
Improvement in range of movement (ROM) in D1 abduction, D1 flexion, D1 extension and D2 abduction for participant CTS01. A) shows superimposed images from week 5 and week 31; B) Change in ROM for the 4 different movements.

## References

Anderson KD (2004) Targeting Recovery: Priorities of the Spinal Cord-Injured Population. Journal of Neurotrauma 21:1371–1383.

Angeli CA, Boakye M, Morton RA, Vogt J, Benton K, Chen Y, Ferreira CK, Harkema SJ (2018) Recovery of Over-Ground Walking after Chronic Motor Complete Spinal Cord Injury. New England Journal of Medicine 379:1244–1250.

Angeli CA, Edgerton VR, Gerasimenko YP, Harkema SJ (2014) Altering spinal cord excitability enables voluntary movements after chronic complete paralysis in humans. Brain 137:1394– 1409 Available at: https://academic.oup.com/brain/article-lookup/doi/10.1093/brain/awu038 [Accessed June 25, 2019].

Benavides FD, Jo HJ, Lundell H, Reggie V, Perez MA (2020) Cortical and Subcortical Effects of Transcutaneous Spinal Cord Stimulation in Humans with Tetraplegia.

Bouton CE, Shaikhouni A, Annetta N v., Bockbrader MA, Friedenberg DA, Nielson DM, Sharma G, Sederberg PB, Glenn BC, Mysiw WJ, Morgan AG, Deogaonkar M, Rezai AR (2016) Restoring cortical control of functional movement in a human with quadriplegia. Nature 533:247–250.

Calvert JS, Manson GA, Grahn PJ, Sayenko DG (2019) Preferential activation of spinal sensorimotor networks via lateralized transcutaneous spinal stimulation in neurologically intact humans. Journal of Neurophysiology 122:2111–2118 Available at: http://www.jn.org [Accessed February 14, 2022].

Capogrosso M et al. (2016) A brain–spine interface alleviating gait deficits after spinal cord injury in primates. Nature 539:284–288 Available at: http://www.nature.com/articles/nature20118 [Accessed June 26, 2019].

Capogrosso M, Wenger N, Raspopovic S, Musienko P, Beauparlant J, Luciani LB, Courtine G, Micera S (2013) A computational model for epidural electrical stimulation of spinal sensorimotor circuits. J Neurosci 33:19326–19340.

Chandrasekaran S, Bickel S, Herrero JL, Kim J won, Markowitz N, Espinal E, Bhagat NA, Ramdeo R, Xu J, Glasser MF, Bouton CE, Mehta AD (2021a) Evoking highly focal percepts in the fingertips through targeted stimulation of sulcal regions of the brain for sensory restoration. Brain Stimulation 14:1184–1196.

Chandrasekaran S, Fifer M, Bickel S, Osborn L, Herrero J, Christie B, Xu J, Murphy RKJ, Singh S, Glasser MF, Collinger JL, Gaunt R, Mehta AD, Schwartz A, Bouton CE (2021b) Historical perspectives, challenges, and future directions of implantable brain-computer interfaces for sensorimotor applications. Bioelectronic Medicine 2021 7:1 7:1–11 Available at: https://link.springer.com/articles/10.1186/s42234-021-00076-6 [Accessed May 17, 2022].

Chandrasekaran S, Nanivadekar AC, McKernan G, Helm ER, Boninger ML, Collinger JL, Gaunt RA, Fisher LE (2020) Sensory restoration by epidural stimulation of the lateral spinal cord in upper-limb amputees. Elife 9:1–26.

Fawcett JW et al. (2006) Guidelines for the conduct of clinical trials for spinal cord injury as developed by the ICCP panel: spontaneous recovery after spinal cord injury and statistical power needed for therapeutic clinical trials. Spinal Cord 2007 45:3 45:190–205 Available at: https://www.nature.com/articles/3102007 [Accessed December 5, 2021].

Fifer MS, McMullen DP, Osborn LE, Thomas TM, Christie BP, Nickl RW, Candrea DN, Pohlmeyer EA, Thompson MC, Anaya MA, Schellekens W, Ramsey NF, Bensmaia SJ, Anderson WS, Wester BA, Crone NE, Celnik PA, Cantarero GL, Tenore F v (2021) Intracortical Somatosensory Stimulation to Elicit Fingertip Sensations in an Individual With Spinal Cord Injury. Neurology:10.1212/WNL.0000000000013173.

Flesher SN, Collinger JL, Foldes ST, Weiss JM, Downey JE, Tyler-Kabara EC, Bensmaia SJ, Schwartz AB, Boninger ML, Gaunt RA (2016) Intracortical microstimulation of human somatosensory cortex. Science Translational Medicine 8.

Freyvert Y, Yong NA, Morikawa E, Zdunowski S, Sarino ME, Gerasimenko Y, Edgerton VR, Lu DC (2018) Engaging cervical spinal circuitry with non-invasive spinal stimulation and buspirone to restore hand function in chronic motor complete patients. Scientific Reports 8:15546 Available at: https://www.nature.com/articles/s41598-018-33123-5.

Friedenberg DA, Schwemmer MA, Landgraf AJ, Annetta N v., Bockbrader MA, Bouton CE, Zhang M, Rezai AR, Mysiw WJ, Bresler HS, Sharma G (2017) Neuroprosthetic-enabled control of graded arm muscle contraction in a paralyzed human. Scientific Reports 2017 7:x1 7:1–10 Available at: https://www.nature.com/articles/s41598-017-08120-9 [Accessed May 17, 2022].

Gad P, Lee S, Terrafranca N, Zhong H, Turner A, Gerasimenko Y, Edgerton VR (2018) Non-invasive activation of cervical spinal networks after severe paralysis. Journal of Neurotrauma 35:2145–2158 Available at: https://www.ncbi.nlm.nih.gov/pmc/articles/PMC6119225/pdf/neu.2017.5461.pdf.

Gerasimenko Y, Gorodnichev R, Moshonkina T, Sayenko D, Gad P, Reggie Edgerton V (2015) Transcutaneous electrical spinal-cord stimulation in humans. Annals of Physical and Rehabilitation Medicine 58:225–231.

Gill ML, Grahn PJ, Calvert JS, Linde MB, Lavrov IA, Strommen JA, Beck LA, Sayenko DG, van Straaten MG, Drubach DI, Veith DD, Thoreson AR, Lopez C, Gerasimenko YP, Edgerton VR, Lee KH, Zhao KD (2018) Neuromodulation of lumbosacral spinal networks enables independent stepping after complete paraplegia. Nature Medicine 2018 24:11 24:1677– 1682 Available at: https://www.nature.com/articles/s41591-018-0175-7 [Accessed February 7, 2022].

Greiner N, Barra B, Schiavone G, James N, Fallegger F, Borgognon S, Lacour S, Bloch J, Courtine G, Capogrosso M (2020) Recruitment of Upper-Limb Motoneurons with Epidural Electrical Stimulation of the Primate Cervical Spinal Cord. bioRxiv:2020.02.17.952796 Available at: https://www.biorxiv.org/content/10.1101/2020.02.17.952796v1.

Guiho T, Baker SN, Jackson A (2021) Epidural and transcutaneous spinal cord stimulation facilitates descending inputs to upper-limb motoneurons in monkeys. Journal of Neural Engineering 18:046011.

Harkema S, Gerasimenko Y, Hodes J, Burdick J, Angeli C, Chen Y, Ferreira C, Willhite A, Rejc E, Grossman RG, Edgerton VR (2011) Effect of epidural stimulation of the lumbosacral spinal cord on voluntary movement, standing, and assisted stepping after motor complete paraplegia: A case study. The Lancet 377:1938–1947 Available at: http://dx.doi.org/10.1016/S0140-6736(11)60547-3 [Accessed June 25, 2019].

Hofstoetter US, Freundl B, Binder H, Minassian K (2018) Common neural structures activated by epidural and transcutaneous lumbar spinal cord stimulation: Elicitation of posterior root-muscle reflexes. PLoS One 13 Available at: https://pubmed.ncbi.nlm.nih.gov/29381748/ [Accessed December 21, 2021].

Inanici F, Brighton LN, Samejima S, Hofstetter CP, Moritz CT (2021) Transcutaneous Spinal Cord Stimulation Restores Hand and Arm Function after Spinal Cord Injury. IEEE Transactions on Neural Systems and Rehabilitation Engineering 29:310–319.

James ND, McMahon SB, Field-Fote EC, Bradbury EJ (2018) Neuromodulation in the restoration of function after spinal cord injury. Lancet Neurol 17:905–917.

Krenn M, Hofstoetter US, Danner SM, Minassian K, Mayr W (2015) Multi-Electrode Array for Transcutaneous Lumbar Posterior Root Stimulation. Artificial Organs 39:834–840 Available at: https://onlinelibrary.wiley.com/doi/full/10.1111/aor.12616 [Accessed February 14, 2022].

Ladenbauer J, Minassian K, Hofstoetter US, Dimitrijevic MR, Rattay F (2010) Stimulation of the human lumbar spinal cord with implanted and surface electrodes: A computer simulation study. IEEE Transactions on Neural Systems and Rehabilitation Engineering 18:637–645.

Lopez L, Sdrulla AD (2021) Success with dorsal root entry zone lesioning after a failed trial of spinal cord stimulation in a patient with pain due to brachial plexus avulsion. PAIN Reports 6:e973.

Lu DC, Edgerton VR, Modaber M, Auyong N, Morikawa E, Zdunowski S, Sarino ME, Sarrafzadeh M, Nuwer MR, Roy RR, Gerasimenko Y (2016) Engaging Cervical Spinal Cord Networks to Reenable Volitional Control of Hand Function in Tetraplegic Patients. Neurorehabilitation and Neural Repair 30:951–962 Available at: https://journals.sagepub.com/doi/full/10.1177/1545968316644344 [Accessed December 6, 2021].

NSCISC, University of Alabama at Birmingham (2021) National Spinal Cord Injury Statistical Center, Facts and Figures at a Glance. Available at: https://www.msktc.org/sci/model-system-centers. [Accessed December 15, 2021].

Rattay F, Minassian K, Dimitrijevic MR (2000) Epidural electrical stimulation of posterior structures of the human lumbosacral cord: 2. quantitative analysis by computer modeling. Spinal Cord 38:473–489.

Rejc E, Angeli CA, Atkinson D, Harkema SJ (2017) Motor recovery after activity-based training with spinal cord epidural stimulation in a chronic motor complete paraplegic. Scientific Reports 7:1–12.

Rowald A et al. (2022) Activity-dependent spinal cord neuromodulation rapidly restores trunk and leg motor functions after complete paralysis. Nature Medicine 2022 28:2 28:260–271 Available at: https://www.nature.com/articles/s41591-021-01663-5 [Accessed March 22, 2022].

Sayenko DG, Atkinson DA, Dy CJ, Gurley KM, Smith VL, Angeli C, Harkema SJ, Edgerton VR, Gerasimenko YP (2015) Spinal segment-specific transcutaneous stimulation differentially shapes activation pattern among motor pools in humans. Journal of Applied Physiology 118:1364–1374.

Sayenko DG, Rath M, Ferguson AR, Burdick JW, Havton LA, Edgerton VR, Gerasimenko YP (2019) Self-Assisted Standing Enabled by Non-Invasive Spinal Stimulation after Spinal Cord Injury. Journal of Neurotrauma 36:1435–1450.

Seáñez I, Capogrosso M (2021) Motor improvements enabled by spinal cord stimulation combined with physical training after spinal cord injury: review of experimental evidence in animals and humans. Bioelectronic Medicine 2021 7:1 7:1–13 Available at: https://bioelecmed.biomedcentral.com/articles/10.1186/s42234-021-00077-5 [Accessed February 14, 2022].

Sharma G, Friedenberg DA, Annetta N, Glenn B, Bockbrader M, Majstorovic C, Domas S, Mysiw WJ, Rezai A, Bouton C (2016) Using an artificial neural bypass to restore cortical control of rhythmic movements in a human with quadriplegia. Scientific Reports 6.

Taccola G, Barber S, Horner PJ, Bazo HAC, Sayenko D (2020) Complications of epidural spinal stimulation: lessons from the past and alternatives for the future. Spinal Cord 58:1049– 1059.

van den Brand R, Heutschi J, Barraud Q, DiGiovanna J, Bartholdi K, Huerlimann M, Friedli L, Vollenweider I, Moraud EM, Duis S, Dominici N, Micera S, Musienko P, Courtine G (2012) Restoring voluntary control of locomotion after paralyzing spinal cord injury. Science (1979) 336:1182–1185 Available at: https://www.science.org/doi/abs/10.1126/science.1217416 [Accessed December 6, 2021].

Wagner FB et al. (2018) Targeted neurotechnology restores walking in humans with spinal cord injury. Nature 2018 563:7729 563:65–71 Available at: https://www.nature.com/articles/s41586-018-0649-2 [Accessed December 5, 2021].

Zhang F, Momeni K, Ramanujam A, Ravi M, Carnahan J, Kirshblum S, Forrest GF (2020) Cervical Spinal Cord Transcutaneous Stimulation Improves Upper Extremity and Hand Function in People with Complete Tetraplegia: A Case Study. IEEE Transactions on Neural Systems and Rehabilitation Engineering 28:3167–3174.

